# Association of nocebo hyperalgesia and basic somatosensory characteristics in a large cohort

**DOI:** 10.1101/2020.09.08.20190728

**Authors:** Mari Hanna Feldhaus, Björn Horing, Christian Sprenger, Christian Büchel

## Abstract

Medical outcomes are strongly affected by placebo and nocebo effects. Prediction of who responds to such expectation effects has proven to be challenging. Most recent approaches to prediction have focused on placebo effects in the context of previous treatment experiences and expectancies, or personality traits. However, a recent model has suggested that basic somatosensory characteristics play an important role in expectation responses. Consequently, this study investigated not only the role of psychological variables, but also of basic somatosensory characteristics. In this study, 624 participants underwent a placebo and nocebo heat pain paradigm. Additionally, individual psychological and somatosensory characteristics were assessed. While no significant correlations for placebo responses were identified, nocebo responses were correlated with personality traits (e.g. neuroticism) and somatosensory characteristics (e.g. thermal pain threshold). Importantly, the correlations between somatosensory characteristics and nocebo responses were among the strongest. This study shows that apart from personality traits, basic somatosensory characteristics play an important role in individual nocebo responses, in agreement with the novel idea that nocebo responses result from the integration of top-down expectation and bottom-up sensory information.

## Introduction

Pain is a multi-faceted phenomenon, with a large prevalence as a clinical symptom^1^, substantial impact on quality of life and high societal costs^2^. As with most subjective clinical symptoms, expectations substantially shape pain. Implicit and explicit expectations can arise from sources as diverse as previous symptom experiences, verbal information or suggestions, or social observation^3^. In the case of pain, this has been demonstrated by effects on subjective ratings^4^, in pain-related neuronal changes in the brain and in the spinal cord^5,6^. Expectation-based positive treatment effects are commonly referred to as placebo effects, whereas negative effects (such as worsening of the outcome or the occurrence of side effects) are referred to as nocebo effects^3,7^. They substantially influence treatment success^8^ and the occurrence of adverse side effects^9^. Consequently, placebo and nocebo effects significantly shape patients’ everyday experiences with medical treatments making them an integral part of the treatment itself.

Even for established treatments, experiences based on conditioning processes as well as explicit expectations have been shown to substantially contribute to therapeutic outcome^9^. For example, the disclosure of possible side effects of a treatment already generates a nocebo expectation which can have negative consequences on the treatment outcome^10^. However, the impact of placebo and especially nocebo effects is ostensibly underestimated^4^. Consequently, studying the essential psychophysiological mechanisms and individual psychological and physiological foundations represents a major scientific and clinical objective. Moreover, the identification of key parameters determining placebo and nocebo responses can pave the way for individualized treatment optimization and minimizing expectation-based responses in clinical trials^4^.

Overarching concepts like the biopsychosocial model^11^ have been applied to expectation effects^12,13^ and emphasize the role of multiple classes of characteristics in their genesis. For example, the experience of pain would not only be affected by a patient’s illness itself and individual factors (e.g. personality, coping style), but also by physiological characteristics (e.g. nervous system makeup) and the social context (e.g. reinforcement or rejection). In the scope of this experimental study, both biological and psychological determinants were considered.

To date, possible correlations of placebo and nocebo effects with individual characteristics have been investigated mainly in the psychological domain, especially with a focus on personality traits. For example, previous studies have identified dispositional optimism^14,15^, low state anxiety^15^, openness^16^, interoceptive awareness^16^, extraversion^15^, and reward sensitivity associated traits such as novelty seeking and behavioral drive^17^ as predictors for placebo effect responders. Regarding predictors for nocebo responses, a relationship with dispositional pessimism^14^, and neuroticism^18^ has been reported. In general, placebo effects have received far more attention than nocebo effects^7^, including recent studies involving large cohorts^19,20^. However, the importance of nocebo effects for quality of life, medication adherence and ultimately treatment success are being increasingly acknowledged^21^. In addition to personality traits, other features probably also play a role in the individual’s disposition for placebo and nocebo effects, such as cognitive factors like attention, which have been shown to interfere with pain processing^22,23^. In a novel framework, we highlighted the importance of the integration of expectation and sensory information to form a pain percept in the context of expectancy effects^24^. This notion has received conceptual^25^ and experimental support^26^ and implies that individual somatosensory characteristics such as pain thresholds also influence placebo and nocebo effects.

Based on these premises, we conducted a study investigating placebo and nocebo effects on pain, and their relation to psychological traits, cognitive factors, and more importantly, somatosensory characteristics in a large sample of healthy participants.

## Results

The aim of this study was to identify associations of placebo and nocebo effects with individual characteristics. In a first step, we tested for placebo and nocebo responses using paired t-tests and repeated measures ANOVA. The pairwise t-tests comparing sham treatment and control conditions revealed significant effects for placebo expectation (1.1% mean pain relief [95% CI, 0.1 to 2.0], *p* = 0.028), placebo conditioning (3.3% mean pain relief [95% CI, 2.4 to 4.3], *p* < 0.001), nocebo expectation (7.2% mean pain increase [95% CI, 5.9 to 8.6], *p* < 0.001), and nocebo conditioning (11.9% mean pain increase [95% CI, 10.4 to 13.4], *p* < 0.001). The repeated measures ANOVA revealed that nocebo effects were significantly stronger than placebo effects (*F*(1,2400) = 30.7, *p* < 0.001), independent of conditioning. Conditioning enhanced placebo and nocebo effects compared to manipulating expectation alone (*F*(1,2400) = 7.6, *p* = 0.006). Figure 1A illustrates the main effects.

To identify associations between placebo and nocebo effects and individual characteristics, a regression analysis method that performs both variable selection and regularization (LASSO) was employed in order to enhance the prediction accuracy and thereby increasing interpretability of the results. LASSO was iterated 1000 times to ensure stable results and to estimate standard deviations of coefficients. Figure 1B illustrates the mean coefficients of the ten strongest predictors with standard deviations. No associations with individual characteristics were determined for placebo expectation. For all other modalities, several predictors were identified: in placebo conditioning several principal components were selected inconsistently (35% of iterations), while in both nocebo modalities, several principal components were selected very consistently (close to 100% of iterations; see Supplement Fig. S1 for detailed selection rates). For placebo conditioning 0.2% of the variance was explained (*R*^2^ = 0.002 [SD 0.003]). In contrast, for nocebo expectancy 8.8% of variance was explained (*R*^2^ = 0.088 [SD 0.002]) and for nocebo conditioning more than 10% of the variance was explained (*R*^2^ = 0.101 [SD 0.002]) by the model.

Three of the six quantitative sensory test (QST) principal components showed strong associations with nocebo responses: Higher thermal pain thresholds (QST1) and higher mechanical pain thresholds (QST2) were associated with higher nocebo responses in both nocebo modalities. Furthermore, regarding nocebo expectation, lower detection thresholds (QST3) were associated with higher nocebo responses.

In addition to QST variables, sex was related to both nocebo modalities: Women showed larger nocebo responses than men. With regard to personality traits, higher action orientation during successful performance of activities (ACS2) was associated with higher nocebo responses in both nocebo modalities. Higher neuroticism scores (EPQ2) were also associated with higher nocebo responses in both nocebo modalities and with lower placebo responses in placebo conditioning. Moreover, higher cooperativeness and reward dependence (TCI2) and lower extraversion (EPQ3) were associated with higher nocebo responses in nocebo conditioning.

Working memory performance (n-back1) correlated with nocebo responses in both nocebo modalities, i.e. participants performing better in the more difficult task were more susceptible to nocebo responses. Moreover, higher distractibility from pain due to working memory load (n-back hypoalgesia) was associated with higher nocebo responses. For individual nocebo responses, control ratings and treatment ratings for the ten highest correlated principal components, see Supplement Figure S2.

**Fig. 1:**
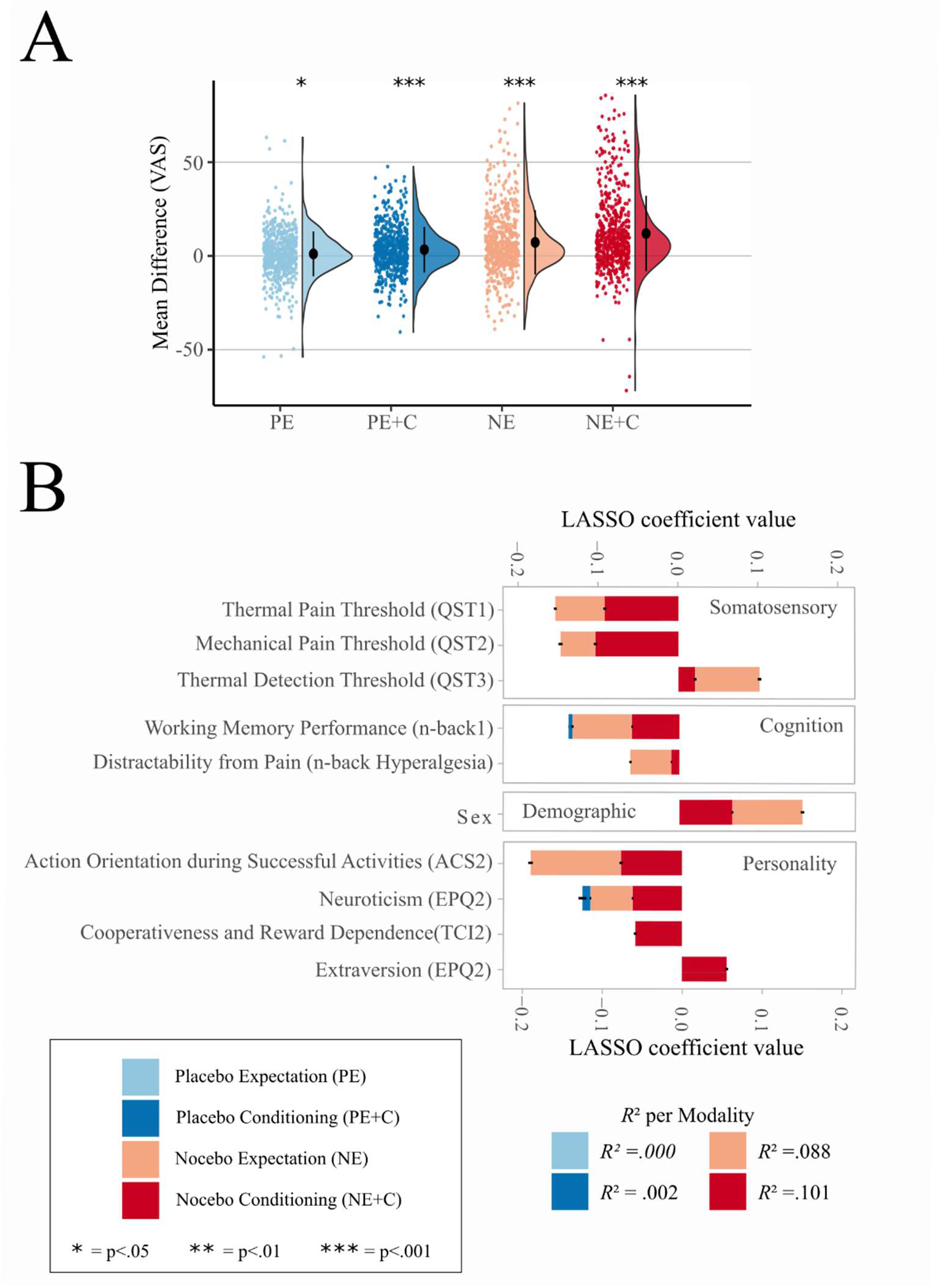
Main effects and LASSO analysis. (A) Group mean of placebo and nocebo effects (difference between sham treatment and control). The colored dots on the left represent raw jittered data of individual participants. The half-violin plots on the right represent the distribution of the data. The black dot represents the group mean and the error bars represent the standard deviation. The figure is color-coded for: Placebo Expectation (PE), Placebo Conditioning (PE+C), Nocebo Expectation (NE), Nocebo Conditioning (NE+C). (B) LASSO results. The stacked bar plots display the ten best LASSO coefficients for each modality with standard deviations of 1000 iterations. Variables are ordered by variable subgroup and within these subgroups ranked by summed overall coefficients.

## Discussion

This large sample study investigated associations of a wide set of individual characteristics with placebo and nocebo responses in pain. We replicated correlations between nocebo responses and certain previously reported personality traits such as neuroticism^18^, extraversion^33^ and reward processing^17^. Furthermore, we observed a connection of distraction-induced hypoalgesia and nocebo effects. More importantly, we observed a significant correlation between nocebo effects and basic somatosensory traits as revealed by quantitative sensory testing. The latter result indicates that individual somatosensory characteristics are as important as psychological traits in determining expectation-dependent pain modulation.

Treatment experience as implemented through conditioning enhanced both placebo and nocebo responses. This result replicates meta-analyses^34,35^, highlighting the importance of personal experience in the clinical setting^19,36^. In clinical practice, patients usually experience repeated treatments and based on the success of previous treatments, form a conscious or unconscious expectation for the current treatment^36^.

Concerning the associations of individual characteristics with placebo and nocebo responses, mainly associations between nocebo responses and individual characteristics were identified. Several correlations with individual characteristics were significant for both nocebo effects (nocebo expectancy and nocebo expectancy plus conditioning). Most importantly, our data showed that basic sensory traits are associated with nocebo responses. Nocebo responses decreased with thermal detection thresholds (QST3), indicating that individuals with a more sensitive thermoception are more susceptible to negative expectations. A possible mechanism is attentiveness: Individuals with lower thermal detection threshold might have been more attentive during threshold estimation and therefore responded faster (yielding a lower threshold). The same individuals might have been also more attentive during nocebo instructions and therefore developed greater expectations and experienced greater nocebo effects.

Furthermore, nocebo responses were positively correlated with thermal pain thresholds (QST1) and mechanical pain thresholds (QST2) revealing that individuals with a higher pain threshold are influenced more strongly by negative treatment expectations. Under the assumption that individuals with a higher pain threshold need more nociceptive input to experience pain, the larger nocebo responses could be related to less precise processing of nociceptive information. A previously introduced Bayesian framework of how expectations can influence pain^24,25^ considers pain perception as the integration of prior knowledge (e.g. pain expectations and experiences) and incoming nociceptive information, which forms and updates the actual pain perception. According to this framework, the precision of nociception determines its relative contribution to actual pain perception: If the precision of nociceptive information is lower compared to the precision of the expectation (prior), the expectation gains more weight in the final perception. In other words: less precise nociception gives room for a stronger effect of expectations and thus a stronger nocebo response (see Supplement Fig. S3 for an illustration). This finding is important, as it can help clinicians to reduce possible side effects by assessing the somatosensory characteristics in patients. Furthermore, the knowledge of these predictors allows to minimize these effects in randomized control trials, increasing assay sensitivity.

Research to identify associations of personality traits with placebo and nocebo responses reaches back decades^37^. However, it has been criticized based on whether context dependency allows unambiguous results^38^. Only replications across different contexts (e.g., experiments) allow to conclude some degree of context independence. Our data replicated several personality traits which have been associated with nocebo effects before, namely neuroticism^18^ and reward sensitivity^17^, making contextual effects less likely. In particular, our data shows that higher neuroticism (EPQ2) predicted nocebo responses, which matches the finding of Davis and colleagues^18^ that higher neuroticism scores are related to greater nocebo responses. We did not observe the previously reported association of placebo and nocebo responses with dispositional optimism^14,15^. Nevertheless, neuroticism is often moderately correlated with optimism^39^, which could explain why some studies identify optimism and others neuroticism as predictors. Moreover, a correlation of lower extraversion and conditioned nocebo responses was identified, which is consistent with other nocebo and placebo findings^33^.

Furthermore, nocebo responses were partially predicted by cooperativeness and reward dependence (TCI2), and action orientation during successful performance of activities (ACS2). The latter trait is related to self-regulation which has been identified as an important mechanism in expectation-based pain modulation^16^. Importantly, both traits are connected to reward: cooperativeness and reward dependence entail social reward, whereas action orientation during successful performance of activities describes being absorbed into an action and therefore the intrinsic reward of an action itself. These results support the notion by Schweinhardt and colleagues^17^, who postulated a relationship between placebo responsiveness and reward sensitivity associated personality traits.

The correlation of sex and nocebo responses was among the strongest, with women showing greater nocebo responses than men. This is in line with a meta-analysis covering six experimental nocebo studies^40^. However, it should be noted that in our study and in at least four out of six studies of the meta-analysis experimenters were all female. As individual pain experience is influenced by the experimenter’s sex^41^, future studies should further investigate the interaction of the participant’s and the experimenter’s sex on placebo and nocebo effects. In line with this and with our own results, a recent study demonstrated that participants reported more side effects if a person from the same gender was present^42^.

Nocebo responses were correlated with cognitive load induced hypoalgesia: Participants who experienced less pain while involved in a demanding working memory task showed greater nocebo responses. While Buhle and colleagues^22^ found evidence for a dissociation of distraction and placebo expectation as pain modulators, our data suggests that at least for nocebo effects these two processes might be associated with each other. Nocebo responses were also predicted by better performances in the more difficult 2-back working memory task. Therefore, individuals who performed better and experienced less pain during the task, showed greater nocebo responses in the experiment. A possible mechanism is that these individuals might be generally more prone to externally initiated pain modulation.

The strength of this study is the large number of analyzed participants as well as the comprehensive set of characteristics that were investigated for correlations with placebo and nocebo effects. However, this study also has several limitations. First, to prevent unnecessary variance between participants, we used a fixed experimental order for all participants. Therefore, carry-over effects can occur. Second, the average placebo effects of 1 to 3 VAS points observed in this study are small, yet within the range of previously reported studies^34,35^. Nevertheless, the methodology employed here has repeatedly proven successful in inducing placebo effects in the past^5,26,43^.. Nevertheless, other studies of comparable sample size have reported larger placebo effects^19,20^. It is therefore possible that the correlation analysis for nocebo responses was more successful because of the higher effect sizes. Third, the assumptions regarding the association of trait variables and outcomes are comparatively basic, and we did not consider more complex psychological process models recently applied to expectation effect research, such as the Elaboration Likelihood Model of persuasion^44^. Relatedly, expectations themselves were not assessed despite conceptual merit^45^, while empirical merit is more ambiguous^19,46^. Here, we opted against querying expectation ratings due to the risk of amplifying demand characteristics and carry-over effects inherent in a cross-over design^47^.

Nocebo mechanisms are an important cause for unspecific side effects. Therefore, it is very important to pinpoint individual characteristics that are associated with nocebo effects and help individualize treatment options. This large study replicated certain personality traits and more importantly, identified additional non-psychological, somatosensory characteristics as predictors for nocebo effects. Hence, conducting selective quantitative assessments of sensory systems in addition to more traditional psychological assessments could help to identify likely nocebo responders in order to improve accuracy in clinical trials and optimize patient treatment. Moreover, the identification of somatosensory characteristics as nocebo predictors highlights the importance of individual psychophysical parameters in a process that has previously been regarded as exclusively psychological in nature.

## Material and Methods

### Study design

In this experimental study, a cross-over design was employed to test placebo and nocebo effects in the form of heat pain hypoalgesia or hyperalgesia. These primary outcomes were used to identify psychological and somatosensory characteristics. Data collection was performed at the University Medical Center Hamburg-Eppendorf. The study was approved by the Ethics Committee of the Medical Board Hamburg, Germany.

### Participants

The sample consisted of 720 healthy participants assessed exclusively for this project. The data was collected from March 2013 to September 2016. Participants gave written informed consent. All participants were fluent German speakers and reported no acute or chronic diseases, no pain medication intake during the last 24 hours, and no damaged skin on either forearm. All participants underwent the same protocol (described in detail in the next paragraphs): They underwent an established placebo and nocebo paradigm^5,6^, filled out a set of questionnaires, completed a quantitative sensory testing (QST)^27^ and performed a working memory paradigm with co-occurring pain stimuli^23^. The experiment lasted for seven hours with a one-hour lunch break. Two participants were investigated at the same time supervised by the experimenter. Participants received €100 as a compensation for their attendance. All participants were debriefed after the experiment and given the opportunity to withdraw their data. No participant withdrew their data. The experiment was performed by two female study psychologists. Both received a 2-day formal training in QST. The first experimenter tested 310 participants, the second experimenter tested 410 participants. In all analyses, experimenter was used as a covariate to control for possible experimenter effects. As participants had to complete a high number of questionnaires and tasks at the computer, certain criteria were defined to identify careless responders that should be excluded. To identify these careless responders, the individual data was scanned for conspicuous patterns in the questionnaires^28^, tasks, and the placebo and nocebo paradigm (i.e. giving different answers to very similar questions or giving the same answer for more than 80 times in a row). These criteria led to the exclusion of 96 participants, resulting in a final sample of 624 participants (373 female, mean 24.6 [SD 3.6] years, range: 18-35) who were included in the data analysis. For detailed exclusion criteria see Supplement Table S1. Detailed sample characteristics are provided in the Supplement Table S2.

### Pain stimulation

Pain was evoked via a contact heat thermode (ATS-Thermode, Medoc LTD Advanced Medical Systems, Rimat Yishai, Israel). The contact area (30×30mm) was attached to skin sites on the volar forearm. Baseline temperature was set to 32°C and painful stimuli ranged from 42 to 48°C. The heat stimuli duration was 10s with a rise and fall rate of 8°C/s. Figure 2 illustrates an experimental trial as it was used during calibration and test. At first, participants watched a black screen with a white cross. As soon as the white cross turned red, the heat stimuli began. Afterwards participants were asked to rate perceived pain on a visual analog scale (VAS) ranging from 0 (“no pain”) to 100 (“unbearable pain”) using keyboard arrow keys and confirming their rating with enter.

**Fig. 2:**
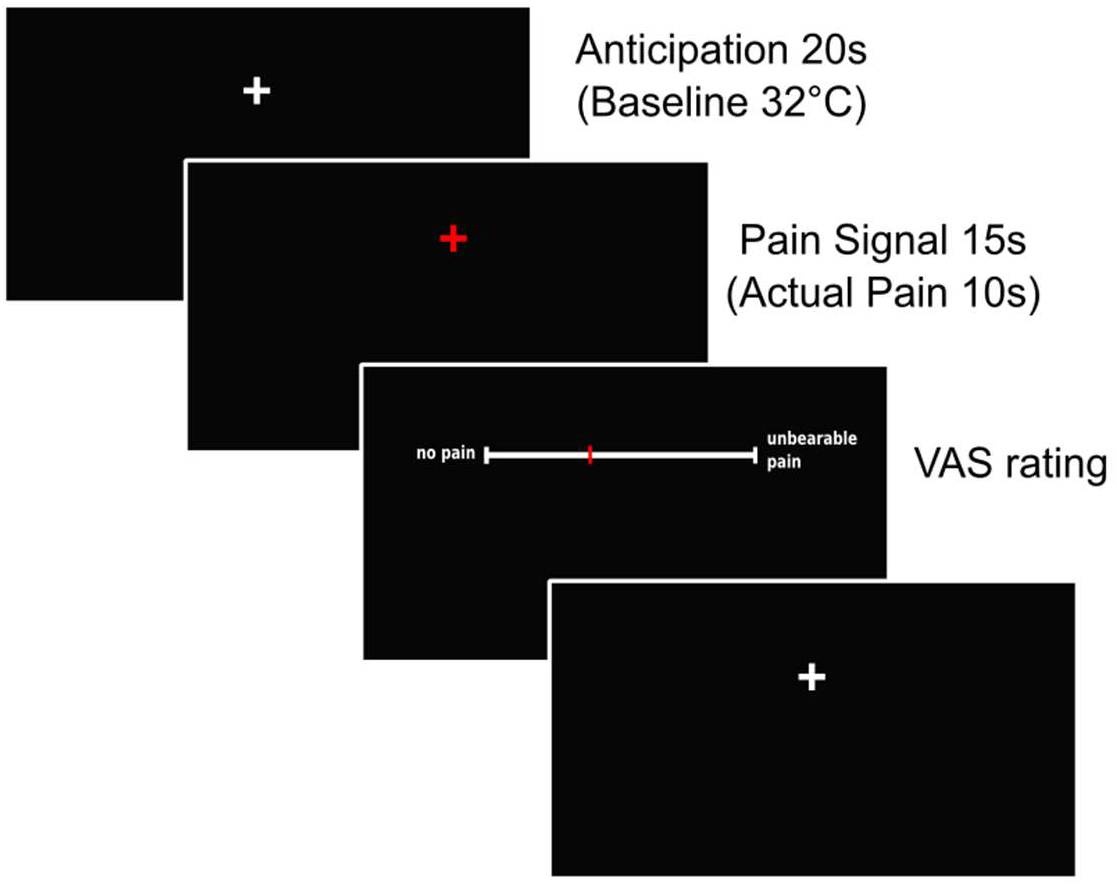
Pain stimulation during one experimental trial. Every experimental trial started with an anticipation phase of 20s during which participants saw a white fixation cross. Afterwards, a red fixation cross appeared indicating the pain stimulation. The red fixation cross was shown for 15s, while the actual pain stimulation lasted for 10s. The pain stimulation was followed by the VAS pain rating.

### Pain calibration

Heat levels were individually calibrated for each participant to achieve the same subjective aversive pain experience across participants. Temperatures were individually calibrated to match *VAS40* and *VAS80* using a stepwise procedure of 6 trials to approach a temperature which matched the chosen VAS rating. The mean of the *VAS40* and *VAS80* temperatures was used as the temperature for *VAS60* stimuli. The calibrated mean temperature (±SD) for *VAS40* was 44.8±1.3°C, for *VAS60* 45.7±1.1°C and for *VAS80* 46.7±1.1°C.

### Experimental Protocol

The following paragraphs will further explain the experimental protocol. The placebo and nocebo paradigm is illustrated in Figure 3A and Figure 3B.

**Fig. 3:**
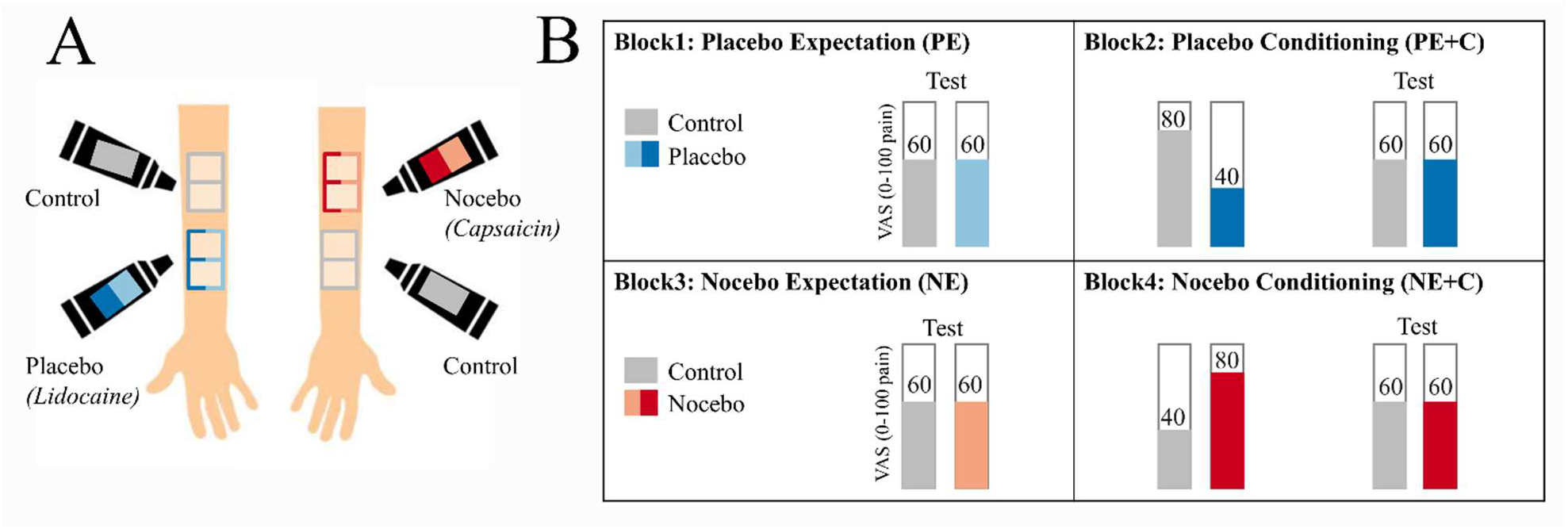
Placebo and nocebo paradigm. (A) Ointments and marked skin areas. For experimental block 1 and 2, a placebo ointment (“Lidocaine”) and a control ointment were introduced. It was explained that “Lidocaine” would exert an analgesic effect. Four skin areas were marked on one forearm of the participants. The skin areas were color-coded for “Lidocaine” and control treatment. For experimental block 1 and the conditioning part of experimental block 2, one control area and one placebo area were used, whereas the second phase (“test”) in experimental block 2 was performed on the remaining two skin areas on the same forearm. For experimental block 3 and 4, the nocebo ointment (“Capsaicin”) was introduced to the participants, as a treatment with a hyperalgesic effect. Four skin areas were marked on the other forearm. For experimental block 3 and the conditioning part experimental block 4, one control area and one nocebo area were used. Afterwards, the second phase (“test”) of experimental block 4 was performed at another nocebo area and control area. The order of the left and right arm and the position of placebo, nocebo and control were counterbalanced across participants. (B) Placebo and nocebo paradigm. In experimental block 1, placebo expectation (PE) effects were tested: Participants were exposed to two times eight heat stimuli, which were calibrated to match 60 on a 0 to 100 VAS (*VAS60*). In experimental block 2, placebo conditioning (PC) effects were tested. Therefore, participants were first conditioned (*VAS80* stimuli for control skin area and *VAS40* for placebo skin area) on the previous skin areas, and then tested again with *VAS60* on the remaining two skin areas. Afterwards participants were introduced to the nocebo treatment and experimental block 3 and 4 were performed in the same manner as experimental block 1 and 2, except that the nocebo skin area was conditioned with *VAS80* and the control skin area was conditioned with *VAS40*, respectively. The overall order of the blocks was fixed for all participants, while the order of treatment (placebo or nocebo) and control was counterbalanced across participants.

### Experimental Introduction

Two participants arrived in the morning and were individually interviewed for any chronic or recent health issues, informed about the pain experiment and gave written informed consent. In the experimental room, participants were seated at two tables, separated by a room divider. The experimenter informed them about the alleged purpose of the study that was to examine associations of pain perception and personality, cognitive abilities, and skin sensitivity in the context of two different treatments. To further explain the two treatments, three different ointments were introduced. The first ointment was introduced as a lidocaine ointment that anesthetizes the skin and therefore decreases pain perception. The second ointment was introduced as a capsaicin ointment that irritates the skin and therefore increases pain perception. The third ointment was introduced as a control ointment that is neutral and has no effect on pain perception. Unbeknown to the participant, all ointments were identical and free of any active ingredient. The first ointment will further be called placebo ointment, the second ointment will further be called nocebo ointment and the third ointment will further be called control ointment. The participants were tested interleaved: When one participant was tested in the placebo- and nocebo paradigm, the other participant was answering questionnaires.

### Block 1 – Placebo Expectation

As illustrated in Figure 3A, four areas were marked on the inner forearm of the first participant. The placebo ointment was applied on two adjacent areas and the control ointment was applied on the other two areas. Left and right forearm as well as upper and lower forearm was randomized across participants. To give the ointment some time to “soak in”, temperatures were now calibrated on another skin area at the same forearm, as explained above. Now, the placebo effect based on pure expectation (by verbal suggestion) was assessed. The participant rated eight medium *VAS60* stimuli on placebo-treated skin area (placebo condition) and eight medium *VAS60* stimuli on a control skin area (control condition).

### Quantitative sensory testing

Next, QST was performed according to the protocol of the German Research Network on Neuropathic Pain^27^ by QST-certified personnel. The following measures were assessed: cold detection threshold, warm detection threshold, cold pain threshold, heat pain threshold, mechanical detection threshold, mechanical pain threshold, dynamic mechanical allodynia, temporal pain summation (“Wind-up”), vibration detection threshold, and pressure pain threshold. For further details see the comprehensive protocol for clinical trials^27^.

### Block 2 – Placebo Expectation plus Conditioning

Now the second part of the placebo paradigm commenced. First, the conditioning procedure was performed: On the previously examined skin areas (in block 1), eight stimuli were applied and rated. Unbeknownst to the participant, the placebo-treated skin area was exposed to eight less painful *VAS40* stimuli to let them experience the effectiveness of the allegedly analgesic ointment, while the control skin area was exposed to eight more painful *VAS80* stimuli. Afterwards, the placebo effect was assessed using the remaining skin areas and applying eight *VAS60* stimuli on both, the placebo-treated and control-treated skin area (identical to block 1).

### Block 3 – Nocebo Expectation

After a one-hour individual lunch break, participants were further introduced to the “pain enhancing” (nocebo) ointment. The nocebo ointment and the control ointment were applied to either two marked areas on the other forearm. The participants answered questionnaires while the ointment “soaked in” for five minutes. Afterwards, they were assessed for the nocebo effect based on pure expectation (by verbal suggestion). The participant rated eight medium *VAS60* stimuli on a nocebo-treated skin area (nocebo condition) and eight medium *VAS60* stimuli on a control skin area (control condition).

### Working memory paradigm

At some point during the alternating experimental tasks [or add the precise point, I don’t think it matters that much], either participant underwent the working memory paradigm. Working memory capacity and distraction induced hypoalgesia were assessed by a working memory paradigm using an n-back task^23^. The task is illustrated in Figure 4. The participants’ task was to concentrate on a stream of letters, which were presented successively in the middle of the screen. For the basic level (“1-back”), the participant had to respond as fast as possible whenever one letter was the same as the letter shown before, as indicated with a red circle in Figure 4 on the left side. For the advanced level (“2-back”), the participant had to react as fast as possible whenever the letter was the same as the one before the last letter, as indicated with a red circle in Figure 4 on the right side. One block consisted of 15 successively presented letters. While the participant was involved in the task, a heat pain stimulus calibrated to match *VAS60* was applied at the participants forearm for each block and afterwards rated on a 0 to 100 VAS scale. Hit rates, false alarm rates, error rates, reaction times and VAS ratings were recorded. We analyzed the difference scores of “2-back” minus “1-back”, as this contrasts high working memory load to low working memory load.

**Fig. 4:**
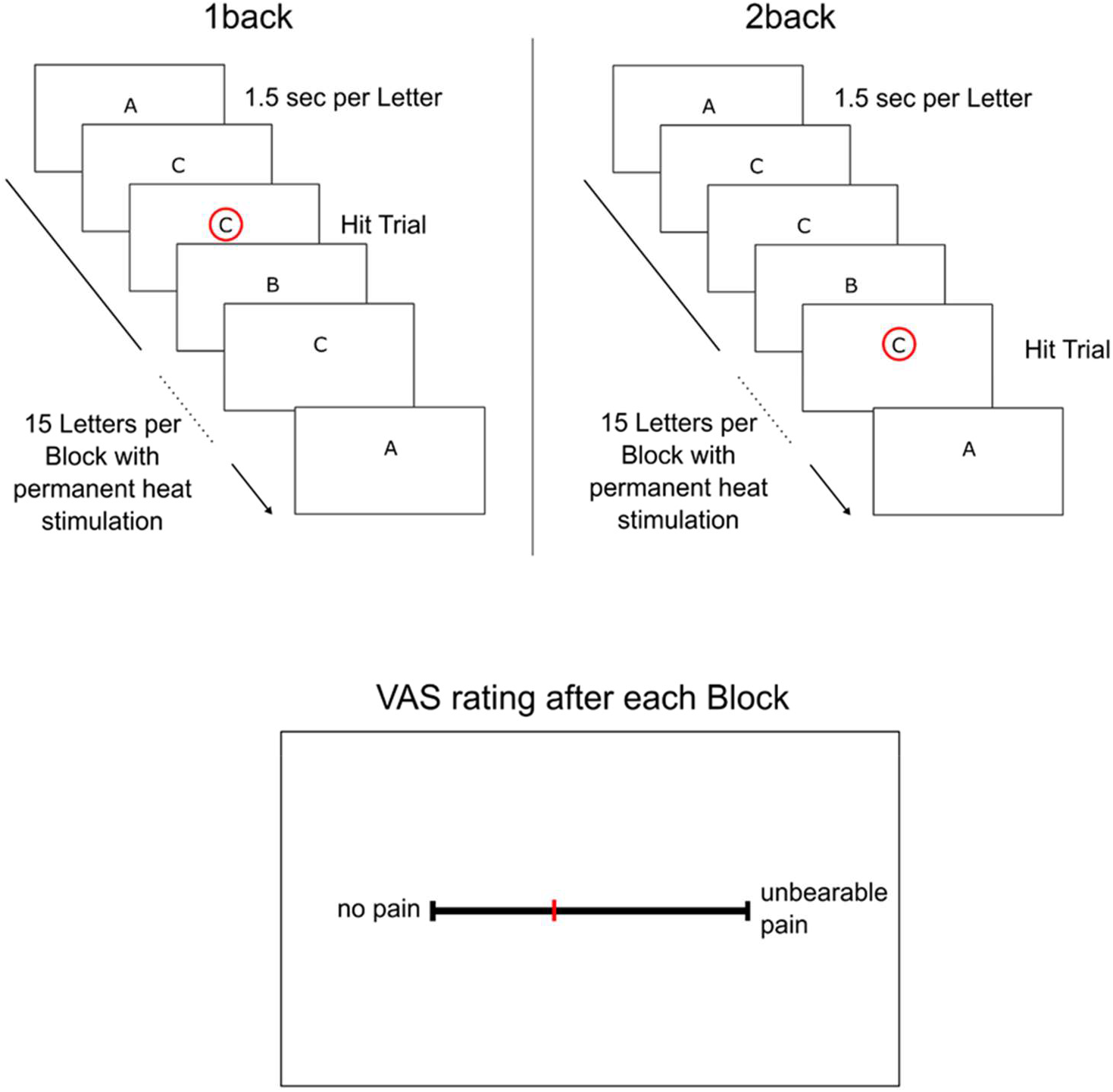
Working Memory Paradigm „n-back”. The upper left part illustrates the easier version of the working memory paradigm (“1-back”). The participant watched a stream of letters and had to respond when the same letter is presented two times in a row, as indicated by the red circle. The upper right part illustrates the more difficult version of the working memory paradigm (“2-back”). The participant watched a stream of letters and had to respond when the letter was the same as the one before the last letter, as indicated by the red circle. While the participant was involved in the task, a heat pain was applied on the forearm, and the participants rated their heat pain experience after each block on a visual analogue scale.

### Block 4 – Nocebo Expectation plus Conditioning

Now the second part of the nocebo paradigm commenced As in block 2, an additional conditioning was introduced: On the previously examined skin areas (in block 3), eight stimuli were applied and rated. Unbeknown to the participant who believed that the temperatures remained unchanged, the nocebo-treated skin area was exposed to eight more painful *VAS80* stimuli to let them experience the effectiveness of the allegedly sensitizing ointment, while the control skin area was exposed to eight *VAS40* stimuli. Afterwards, the nocebo effect was assessed using different skin areas and applying eight *VAS60* stimuli on both, the nocebo-treated and control-treated skin area (identical to block 3). The difference of these ratings was considered as the nocebo effect base on expectation plus conditioning. After the nocebo paradigm was completed participants were fully informed about the actual purpose of the study.

### Questionnaires and additional secondary assessments

Questionnaires were answered whenever the participant was not directly involved in the placebo/nocebo paradigm, QST, or the working memory paradigm. The following questionnaires were assessed: Action Control Scale 90 (ACS-90), Anxiety Sensitivity-Index3 (ASI), Beliefs about Medicines Questionnaires (BMQ), Cognitive Emotion Regulation Questionnaire (CERQ), Center for Epidemiologic Studies Depression Scale (CES-D-Scale), Defensive Pessimism Questionnaire (DPQ), Emotion Regulation Questionnaire (ERQ), German Extended Personal Attributes Questionnaire (GEPAQ), General Competence Expectancy Test (GKE), Internality, Powerful Other and Chance Scale (IPC), Life-Orientation-Test (LOT), Pain Catastrophizing Scale (PCS), Pain Vigilance and Awareness Questionnaire (PVAQ), Symptom Checklist 90 (SCL 90), Social Desirability Scale-17 (SDS-17), State Trait Anxiety Inventory (STAI), Temperament Character Inventory (TCI). Additionally, sex, age, and body mass index (BMI) were assessed.

### Placebo and Nocebo Effect Estimation

Placebo and nocebo were defined as the difference in pain ratings between control ratings and (sham) treatment ratings. Placebo effects were defined as control ratings minus placebo ratings (for identical *VAS60* stimuli), while nocebo effects were defined as nocebo ratings minus control ratings, leading to positively coded effects in placebo and nocebo blocks. This within-participant design allowed for an estimation of placebo and nocebo effects for each participant, which enabled us to identify associations of placebo and nocebo effects with individual traits.

Placebo and nocebo effects are expectancy driven. These expectations can be shaped by verbal suggestions and conditioning, whereas conditioning seems to strengthen verbally suggested expectancy^29–31^. Consequently, we tested placebo and nocebo effects by pure expectation induced by verbal suggestions and by classical conditioning.

The order of the experimental blocks was identical for all participants, as the aim of this study was the identification of predictors for placebo and nocebo effects. Moreover, because of learning effects, the order of pure expectation effects and expectation effects plus conditioning could not be changed. Although a fixed order confounds the relative difference between placebo and nocebo effects with time of test (morning vs afternoon), it also reduces the variance across participants, and thus can increase the statistical power to identify associations with individual characteristics.

### Statistical analysis

To account for collinearity within domains and increase interpretability, measures were restructured using principal component analyses (R package “psych”, version 1.7.8). These were calculated for QST, the cognitive task and the individual questionnaires (using the questionnaire scales). The number of principal components in each domain was chosen as to explain at least 70% of the total variance. This procedure reduced the number of predictors from 75 to 46. All variables and components were rescaled and centered. Principal components were characterized according to the subscales providing the highest loading. An illustration of all individual loadings of the constructed principal components with the original variables can be found in Supplement Figure S4.

Our experimental aim was to identify associations of placebo and nocebo effects with individual characteristics. To prove a successful induction of placebo and nocebo effects, we tested the placebo and nocebo ratings effects using repeated measures ANOVA and paired sample t-tests.

To examine associations of individual characteristics with placebo and nocebo effects, a regularized regression analysis with variable selection (LASSO; least absolute shrinkage and selection operator)^32^ was employed. LASSO minimizes the residual sum of squares by imposing a penalty (regularization) and therefore reduces correlated coefficient values towards zero^32^, which enhances the prediction accuracy and interpretability of the predictors. This penalty for correlated variables was beneficial to our set of principal components, as they could still be correlate given that the PCA was performed within each questionnaire, QST and working memory paradigm. For an intercorrelation analysis of the principal components see Supplement Figure S5. As a result, LASSO obtained a subset of the original characteristics which has the advantage of higher prediction accuracy and better interpretability compared to a nonregularized regression model^32^. LASSO was performed using the R-package “glmnet”, version 2.0-13. Missing data was imputed in each iteration, and results were cross-validated. Because of the imputation of missing data and the cross-validation, LASSO selection and coefficient estimation could differ depending on tuning parameters. Therefore, the analysis was repeated 1000 times for placebo and nocebo responses, which secured stable results. In all analyses, we controlled for experimenter and absolute calibrated temperature as covariates. All analyses were performed using R version 3.4.2 software.

## Data Availability

The data is stored at University Clinic Hamburg-Eppendorf.

## Statements

## Acknowledgements

We would like to thank Kristina Schmitz (University Medical Center Hamburg-Eppendorf, Dipl. Psych), who was employed as a study psychologist for the time of the data analysis for her contribution to the design and data collection. Also, the authors thank Arvina Grahl, Lea Kampermann, Alexandra Tinnermann, Paul Enck, and Tor Wager for helpful comments.

## Statement of Ethics

The study was approved by the Ethics Committee of the Medical Board Hamburg, Germany (PV4030), and it was conducted in accordance with the World Medical Association Declaration of Helsinki. All participants provided verbal and written consent to participate.

## Additional Information

The authors have no conflicts of interest to declare.

## Funding Sources

This study was financially supported by the European Research Council (grant ERC-2010-AdG_20100407). Björn Horing was supported by the Alexander von Humboldt Foundation. However, the funding agency had no role in design and conduct of the study; collection, management, analysis, and interpretation of the data; preparation, review, or approval of the manuscript; and decision to submit the manuscript for publication.

## Author Contributions Statement

CS and CB contributed to the experimental design. MHF collected the data. MHF analyzed the data with the help of BH and CB. MHF created the figures. MHF wrote this article with the help of BH and CB. MHF had full access to all the data in the study and takes responsibility for the integrity of the data and the accuracy of the data analysis.

## Notes

### Competing Interest Statement

The authors have declared no competing interest.

### Clinical Trial

We would like to mention, that our study is post-registered at the DRKS (DRKS00017036) because at the time when the study was designed the implications for clinicians were not as obvious as they are now. We (and colleagues such as Prof. Kaptchuk at Harvard Medical School) strongly believe that our study showing predisposing factors for nocebo responses in a very large sample is unique and extremely important for all clinicians. Further, we would like to point out that our study has been pre-approved by the local ethics committee with a detailed study protocol.

### Funding Statement

This study was financially supported by the European Research Council (grant ERC-2010-AdG_20100407). Bjoern Horing was supported by the Alexander von Humboldt Foundation. However, the funding agency had no role in design and conduct of the study; collection, management, analysis, and interpretation of the data; preparation, review, or approval of the manuscript; and decision to submit the manuscript for publication.

### Author Declarations

The study was approved by the Ethics Committee of the Medical Board Hamburg, Germany.

## References

1. Vos, T. et al. Global, regional, and national incidence, prevalence, and years lived with disability for 301 acute and chronic diseases and injuries in 188 countries, 1990–2013: a systematic analysis for the Global Burden of Disease Study 2013. The Lancet 386, 743–800 (2015).

2. Manchikanti, L., Singh, V., Kaye, A. D. & Hirsch, J. A. Lessons for Better Pain Management in the Future: Learning from the Past. Pain Ther. (2020) doi:10.1007/s40122-020-00170-8.

3. Colloca, L. & Barsky, A. J. Placebo and Nocebo Effects. N. Engl. J. Med. 382, 554–561 (2020).

4. Evers, A. W. M. et al. Implications of placebo and nocebo effects for clinical practice: expert consensus. Psychother. Psychosom. 87, 204–210 (2018).

5. Eippert, F. et al. Activation of the opioidergic descending pain control system underlies placebo analgesia. Neuron 63, 533–43 (2009).

6. Tinnermann, A., Geuter, S., Sprenger, C., Finsterbusch, J. & Büchel, C. Interactions between brain and spinal cord mediate value effects in nocebo hyperalgesia. Science 358, 105–108 (2017).

7. Schedlowski, M., Enck, P., Rief, W. & Bingel, U. Neuro-Bio-Behavioral Mechanisms of Placebo and Nocebo Responses: Implications for Clinical Trials and Clinical Practice. Pharmacol. Rev. 67, 697–730 (2015).

8. Bingel, U. et al. The effect of treatment expectation on drug efficacy: Imaging the analgesic benefit of the opioid remifentanil. Sci. Transl. Med. 3, 70ra14 (2011).

9. Enck, P., Bingel, U., Schedlowski, M. & Rief, W. The placebo response in medicine: Minimize, maximize or personalize? Nat. Rev. Drug Discov. 12, 191–204 (2013).

10. Flaten, M. A., Simonsen, T. & Olsen, H. Drug-related information generates placebo and nocebo responses that modify the drug response. Psychosom. Med. 61, 250–255 (1999).

11. Engel, G. L. The need for a new medical model: a challenge for biomedicine. Science 196, 129–136 (1977).

12. Blasi, Z. D., Harkness, E., Ernst, E., Georgiou, A. & Kleijnen, J. Influence of context effects on health outcomes: a systematic review. The Lancet 357, 757–762 (2001).

13. Fava, G. A., Guidi, J., Rafanelli, C. & Rickels, K. The Clinical Inadequacy of the Placebo Model and the Development of an Alternative Conceptual Framework. Psychother. Psychosom. 86, 332–340 (2017).

14. Geers, A. L., Kosbab, K., Helfer, S. G., Weiland, P. E. & Wellman, J. A. Further evidence for individual differences in placebo responding: An interactionist perspective. J. Psychosom. Res. 62, 563–570 (2007).

15. Morton, D. L., Watson, A., El-Deredy, W. & Jones, A. K. P. Reproducibility of placebo analgesia: Effect of dispositional optimism. Pain 146, 194–198 (2009).

16. Vachon-Presseau, E. et al. Brain and psychological determinants of placebo pill response in chronic pain patients. Nat. Commun. 9, 3397 (2018).

17. Schweinhardt, P., Seminowicz, D. A., Jaeger, E., Duncan, G. H. & Bushnell, M. C. The anatomy of the mesolimbic reward system: A link between personality and the placebo analgesic response. J. Neurosci. 29, 4882–4887 (2009).

18. Davis, C., Ralevski, E., Kennedy, S. H. & Neitzert, C. The role of personality factors in the reporting of side effect complaints to moclobemide and placebo: A study of healthy male and female volunteers. J. Clin. Psychopharmacol. 15, 347–352 (1995).

19. Colloca, L. et al. Prior Therapeutic Experiences, Not Expectation Ratings, Predict Placebo Effects: An Experimental Study in Chronic Pain and Healthy Participants. Psychother. Psychosom. 1–8 (2020) doi:10.1159/000507400.

20. Wang, Y. et al. Modeling Learning Patterns to Predict Placebo Analgesic Effects in Healthy and Chronic Orofacial Pain Participants. Front. Psychiatry 11, 39 (2020).

21. Pouillon, L. et al. The nocebo effect: a clinical challenge in the era of biosimilars. Expert Rev. Clin. Immunol. 14, 739–749 (2018).

22. Buhle, J. T., Stevens, B. L., Friedman, J. J. & Wager, T. D. Distraction and placebo: two separate routes to pain control. Psychol. Sci. 23, 246–53 (2012).

23. Sprenger, C. et al. Attention modulates spinal cord responses to pain. Curr. Biol. 22, 1019–1022 (2012).

24. Büchel, C., Geuter, S., Sprenger, C. & Eippert, F. Placebo analgesia: A predictive coding perspective. Neuron 81, 1223–1239 (2014).

25. Ongaro, G. & Kaptchuk, T. J. Symptom perception, placebo effects, and the Bayesian brain. Pain 160, 1–4 (2019).

26. Grahl, A., Onat, S. & Büchel, C. The periaqueductal gray and Bayesian integration in placebo analgesia. eLife 7, e32930 (2018).

27. Rolke, R. et al. Quantitative sensory testing: A comprehensive protocol for clinical trials. Eur. J. Pain 10, 77–88 (2006).

28. Meade, A. W. & Craig, S. B. Identifying careless responses in survey data. Psychol. Methods 17, 437 (2012).

29. Colloca, L., Sigaudo, M. & Benedetti, F. The role of learning in nocebo and placebo effects: Pain 136, 211–218 (2008).

30. Klinger, R., Soost, S., Flor, H. & Worm, M. Classical conditioning and expectancy in placebo hypoalgesia: A randomized controlled study in patients with atopic dermatitis and persons with healthy skin: Pain 128, 31–39 (2007).

31. Kirsch, I. et al. Expectancy and conditioning in placebo analgesia: Separate or connected processes? Psychol. Conscious. Theory Res. Pract. 1, 51–59 (2014).

32. Tibshirani, R. Regression shrinkage and selection via the LASSO. J. R. Stat. Soc. Ser. B Methodol. 58, 267–288 (1996).

33. Kelley, J. M. et al. Patient and practitioner influences on the placebo effect in irritable bowel syndrome. Psychosom. Med. 71, 789 (2009).

34. Petersen, G. L. et al. The magnitude of nocebo effects in pain: A meta-analysis. Pain 155, 1426–1434 (2014).

35. Vase, L., Petersen, G. L., Riley, J. L. & Price, D. D. Factors contributing to large analgesic effects in placebo mechanism studies conducted between 2002 and 2007. Pain 145, 36–44 (2009).

36. Kessner, S. et al. The Effect of Treatment History on Therapeutic Outcome: Psychological and Neurobiological Underpinnings. PLoS ONE 9, e109014 (2014).

37. Shapiro, A. K. Factors contributing to the placebo effect. Am. J. Psychother. 18, 73–88 (1964).

38. Koban, L., Rozic, L. & Wager, T. D. Brain predictors of individual differences in placebo responding. in Placebo and Pain 89–102 (Academic Press, 2013).

39. Williams, D. G. Dispositional optimism, neuroticism, and extraversion. Personal. Individ. Differ. 13, 475–477 (1992).

40. Vambheim, S. M. & Flaten, M. A. A systematic review of sex differences in the placebo and the nocebo effect. J. Pain Res. 10, 1831–1839 (2017).

41. Kállai, I., Barke, A. & Voss, U. The effects of experimenter characteristics on pain reports in women and men. Pain 112, 142–147 (2004).

42. Mazzoni, G., Foan, L., Hyland, M. E. & Kirsch, I. The effects of observation and gender on psychogenic symptoms. Health Psychol. 29, 181–185 (2010).

43. Geuter, S. & Büchel, C. Facilitation of pain in the human spinal cord by nocebo treatment. J. Neurosci. 33, 13784–13790 (2013).

44. Geers, A. L., Briñol, P. & Petty, R. E. An Analysis of the Basic Processes of Formation and Change of Placebo Expectations. Rev. Gen. Psychol. 23, 211–229 (2019).

45. Peerdeman, K. J. et al. Relieving patients’ pain with expectation interventions: a meta-analysis. PAIN 157, 1179–1191 (2016).

46. Rhudy, J. L., Güereca, Y. M., Kuhn, B. L., Palit, S. & Flaten, M. A. The Influence of Placebo Analgesia Manipulations on Pain Report, the Nociceptive Flexion Reflex, and Autonomic Responses to Pain. J. Pain 19, 1257–1274 (2018).

47. Horing, B., Weimer, K., Muth, E. R. & Enck, P. Prediction of placebo responses: a systematic review of the literature. Front. Psychol. 5, (2014).

